# Scoping Review of Regulatory Transparency in AI-based Radiology Software: Analysis of PMDA-approved SaMD Products

**DOI:** 10.1101/2025.10.02.25336333

**Authors:** Tomohiro Kikuchi, Shannon L Walston, Hirotaka Takita, Yasuhito Mitsuyama, Rintaro Ito, Masahiro Hashimoto, Takeshi Nakaura, Hiroaki Hyakutake, Sho Kawabe, Harushi Mori, Daiju Ueda

## Abstract

**Background:** The integration of artificial intelligence (AI) in radiology has accelerated globally, with Japan’s Pharmaceuticals and Medical Devices Agency (PMDA) approving numerous AI-based Software as a Medical Device (SaMD) products. However, the transparency and completeness of clinical evidence available to healthcare providers remain unclear.

**Purpose:** To systematically evaluate the availability and transparency of clinical evidence in package inserts of PMDA-approved AI-based radiology SaMD products, identifying gaps that may impact clinical implementation.

**Materials and Methods:** We conducted a systematic review of all PMDA-approved SaMD products as of December 31, 2024. Products were included if they utilized AI technology and were classified for radiology applications. Data extraction focused on product characteristics, study designs, demographic information, and performance metrics.

**Results:** Of 151 approved SaMD products, 40 utilized AI technology, with 20 specifically designed for radiology applications. Critical gaps were identified in demographic reporting, with no products providing complete case demographic data. Performance metrics varied widely, with sensitivity ranging from 67.7% to 100% in standalone studies. Physician-assisted studies consistently demonstrated performance improvements but lacked stratified results by characteristics in all cases.

**Conclusion:** Current package insert requirements provide insufficient transparency for evidence-based clinical implementation of AI-based radiology software. Enhanced regulatory frameworks and industry-led initiatives for comprehensive validation are essential for safe and effective AI deployment in Japanese healthcare.

## Introduction

The rapid advancement of artificial intelligence (AI) technologies has fundamentally transformed the landscape of medical imaging and radiology practice worldwide [1]. In Japan, the Pharmaceuticals and Medical Devices Agency (PMDA) has established itself as a pioneering regulatory body in the approval of AI-based Software as Medical Device (SaMD) products, with a notable concentration in radiology applications[2]. As of 2024, Japan is an early adopter of AI-based medical devices, with a dedicated SaMD review office and dozens of approved products, reflecting its technologically advanced healthcare system and progressive regulatory framework [3].

The integration of AI into clinical radiology practice presents unique challenges that extend beyond traditional medical device implementation. Unlike conventional imaging equipment, AI-based software systems exhibit performance characteristics that can vary significantly based on patient demographics, imaging protocols, and institutional practices [4]. This variability necessitates a higher standard of transparency in clinical evidence to enable healthcare providers to make informed decisions about implementation and to understand the limitations of these technologies in their specific clinical contexts [5].

Package inserts, known as “tenpu-bunsho” in Japanese regulatory terminology, serve as the primary source of product information for clinicians and healthcare institutions [6]. These documents are legally required to contain essential information about the safety, efficacy, and proper use of medical devices [7]. For AI-based SaMD products, package inserts represent a critical communication channel between manufacturers, regulators, and end-users, theoretically providing the evidence base necessary for safe and effective clinical implementation. However, the rapid pace of AI development and deployment has outstripped the evolution of regulatory documentation standards. International studies have highlighted significant variations in the quality and completeness of clinical evidence provided for AI-based medical devices, with particular concerns about the generalizability of performance claims across diverse patient populations [8,9]. The unique characteristics of the Japanese healthcare system, including its demographic composition and clinical practice patterns, make it particularly important to understand how well current regulatory documentation serves the needs of Japanese healthcare providers [10].

Furthermore, the implementation of AI in radiology intersects with Japan’s evolving framework for medical data governance, including the Next Generation Medical Infrastructure Act and the Personal Information Protection Act [10]. These regulations create both opportunities and challenges for the development and validation of AI systems, requiring specialized expertise to navigate the complex landscape of data privacy, clinical validation, and regulatory compliance.

The objective of this study is to systematically evaluate the transparency and completeness of clinical evidence provided in package inserts for PMDA-approved AI-based radiology SaMD products. By analyzing the availability of demographic data, performance metrics, and study design information, we aim to identify gaps that may impact clinical implementation and to provide recommendations for enhancing regulatory transparency in this rapidly evolving field.

## Materials and Methods

### Data Source and Search Strategy

On 31 May 2025 we downloaded the Excel spreadsheet listing all PMDA-approved SaMD products from the PMDA website [11]. We filtered the data to include only records approved on or before 31 December 2024. Two investigators independently applied these filters, then compared outputs and confirmed complete concordance.

### Inclusion and Exclusion Criteria

Products were included in the analysis if they met the following criteria: (1) approved by PMDA as SaMD, (2) explicitly described as utilizing artificial intelligence, and (3) indicated for use in radiology applications, including but not limited to X-ray, computed tomography (CT), magnetic resonance imaging (MRI), and ultrasound imaging. Products were excluded if they were (1) non-AI software; (2) applications dedicated to endoscopy or laparoscopy; (3) software intended for pathology, other specimen-based laboratory testing, or electrocardiography—applications not directly related to radiology; or (4) consumer-oriented wearable products such as Apple Watch applications. This exclusion criteria ensured focus on traditional radiology imaging modalities.

### Data Extraction Framework

The extraction framework encompassed the following categories: Product characteristics included approval ID, version number, product name and overview, imaging modality, marketing authorization holder, manufacturer, and study design used for regulatory approval. For each product, we documented whether approval was based on standalone performance testing, physician-assisted studies, or both.

Clinical study characteristics were extracted in detail, including total sample size, number of positive cases, patient demographic information (nationality, ethnicity, race, sex, age distribution), imaging equipment vendors, and participating clinical sites. We specifically noted whether demographic breakdowns were provided and whether performance metrics were stratified by these characteristics.

Performance metrics extraction focused on standard diagnostic accuracy measures including sensitivity, specificity, positive predictive value (PPV), negative predictive value (NPV), area under the receiver operating characteristic curve (AUC), and false positive rates. For physician-assisted studies, we additionally extracted information about reader characteristics, including number of readers, specialization, experience levels, and geographic distribution.

### Data Analysis

Descriptive statistics were used to quantify reporting completeness. For every product we recorded the presence (coded 1) or absence (coded 0) of the following transparency items: total case number; demographic breakdowns for nationality/ethnicity/race, sex, and age; vendor-specific information; site-specific information; and performance metrics stratified by any demographic or technical variable. We then tabulated the number and percentage of products that reported each item; no composite score was calculated and products were not ranked.

Performance metrics were summarized using ranges and medians where appropriate. For products reporting multiple performance values (e.g., at different thresholds or for different findings), we documented the manufacturer-designated primary metric. Physician-assisted studies were analyzed separately to assess the impact of AI assistance on diagnostic performance.

## Results

### Product Landscape and Eligibility

The PMDA data listed 151 SaMD approvals. Of these, 111 products were non-AI and excluded (exclusion criteria 1). Among the remaining 40 AI-enabled products, 15 were endoscopy/laparoscopy applications (exclusion criteria 2), two were pathology/laboratory/electrocardiography applications (exclusion criteria 3), and three were consumer-oriented wearable apps (exclusion criteria 4). Consequently, 20 AI-based radiology products met all eligibility criteria and were included in the analysis (Figure 1). Twelve products had undergone at least one package-insert revision, and our analysis evaluated the latest version available as of June 2025.

**Figure 1:**
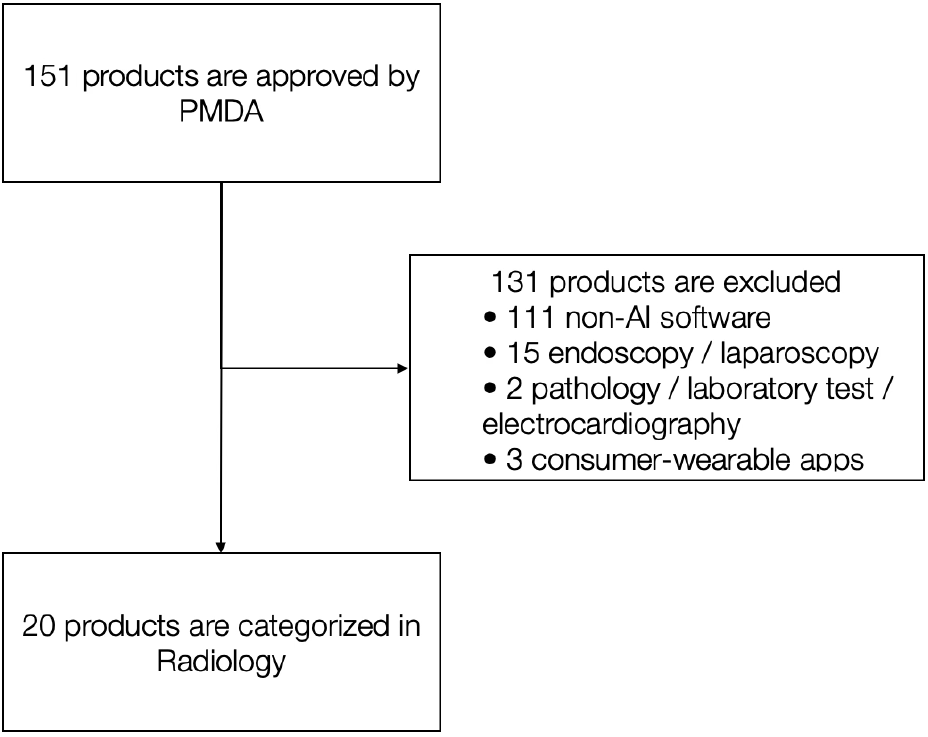
Eligibility. The flowchart illustrates the systematic screening process for identifying eligible AI-SaMDs in radiology. Starting from all SaMDs approved by the PMDA, products were excluded based on pre-defined criteria, resulting in the final cohort for analysis. SaMD, Software as a Medical Device; PMDA, Pharmaceuticals and Medical Devices Agency; AI, Artificial Intelligence.

Table 1 shows a summary of each product. The 20 included radiology AI products demonstrated diverse applications across imaging modalities: CT imaging dominated with eight products [12–19] (40.0%); computed radiography (CR) accounted for seven products [20–26] (35.0%); ultrasound (US) for three products [27–29] (15.0%); MRI for one product [30] (5.0%); and one product (5.0%) had no imaging modality specified [31]. Five products (25.0%) were specifically developed for COVID-19-related imaging findings [13–17].

**Table 1:** Product characteristics.

| Citation | Approved ID | Version | Product overview | Modality | Marketing authorization holder | Manufacturer | Study design for approval |
| --- | --- | --- | --- | --- | --- | --- | --- |
| 12 | 30200BZX0020 2000 | 2nd ed., Apr 2022 | Analyzes chest CT scans to help doctors detect and mark potential lung nodules. | CT | Siemens Healthcare K.K. | Siemens Healthcare GmbH | Standalone + Physician-assisted test |
| 13 | 30200BZX0018 4000 | 2nd ed., Sep 23, 2020 | Analyzes chest CT scans for findings associated with COVID-19 pneumonia, providing reference information. | CT | CES Descartes, Inc. | Beijing Infervision Technology Co.,Ltd. | Standalone test |
| 14 | 30300BZX0014 5000 | 2nd ed., Feb 2024 | Analyzes chest CT scans to show the likelihood of findings seen in COVID-19 pneumonia. | CT | FUJIFILM Corporation | FUJIFILM Corporation | Standalone test |
| 15 | 30200BZX0021 2000 | 1st ed., Jul 2020 | Analyzes chest CT scans to indicate the likelihood of findings related to COVID-19 pneumonia. | CT | MIC Medical Corporation | Alibaba Damo (Hangzhou) Technology Co., Ltd | Standalone test |
| 16 | 30400BZX0012 3000 | 1st ed., Jul 2022 | Analyzes chest CT scans to help identify findings suggestive of COVID-19 pneumonia. | CT | Canon Medical Systems Corporation | Canon Medical Systems Corporation | Standalone test |
| 27 | 30600BZX0015 5000 | 1st ed., Jul 29, 2024 | Analyzes fetal heart ultrasound videos to help doctors assess heart structures and check for abnormalities. | US | Fujitsu Japan Limited | Fujitsu Japan Limited | Standalone + Physician-assisted test |
| 17 | 30300BZX0035 0000 | 5th ed., May 31, 2024 | Analyzes chest CT images to highlight potential findings of COVID-19 pneumonia for doctors. | CT | Fujitsu Japan Limited | Fujitsu Japan Limited | Standalone test |
| 20 | 30600BZX0009 8000 | 1st ed., May 2024 | Analyzes panoramic dental X-rays to help dentists evaluate jawbone thickness and shape. | CR | MEDIA Co., Ltd. | MEDIA Co., Ltd. | Standalone test |
| 30 | 30100BZX0014 2000 | 7th ed., Jun 2027 | Analyzes head MRA scans to help doctors detect potential brain aneurysms. | MR | LPIXEL Inc. | LPIXEL Inc. | Standalone + Physician-assisted test |
| 21 | 30400BZX0028 5000 | 4th ed., May 2024 | Analyzes chest X-rays to help detect and mark potential abnormal shadows, such as nodules or infiltrates. | CR | LPIXEL Inc. | LPIXEL Inc. | Standalone + Physician-assisted test |
| 22 | 30200BZX0026 9000 | 2nd ed., Jun 2024 | Analyzes chest X-rays to help doctors find and mark potential lung nodules. | CR | LPIXEL Inc. | LPIXEL Inc. | Standalone + Physician-assisted test |
| 23 | 30300BZX0027 1000 | 5th ed., Oct 2024 | Analyzes chest X-rays to help detect patterns similar to nodules, masses, and infiltrates. | CR | Konica Minolta, Inc. | Konica Minolta, Inc. | Standalone + Physician-assisted test |
| 24 | 30300BZX0018 8000 | 10th ed., Mar 2025 | Analyzes chest X-rays to detect potential lung nodules, infiltrates, and pneumothorax, showing results as a heatmap. | CR | FUJIFILM Corporation | FUJIFILM Corporation | Standalone + Physician-assisted test |
| 25 | 30300BZX0033 9000 | 1st ed., Jan 2022 | Analyzes chest X-rays to provide a confidence score for findings related to infectious pneumonia. | CR | Doctor-Net Inc. | JLK Inc. | Standalone test |
| 26 | 30500BZX0026 2000 | 1st ed., Dec 2023 | Analyzes chest X-rays to help detect findings suggestive of fibrotic interstitial lung disease. | CR | COSMOTEC Co., Ltd. | COSMOTEC Co., Ltd. | Standalone + Physician-assisted test |
| 28 | 30200BZX0037 9000 | 1st ed., Nov 24, 2020 | Analyzes breast ultrasound images in real-time to help identify and mark potential lesions. | US | CES Descartes, Inc. | TaiHao Medical Inc. (Taiwan) | Standalone + Physician-assisted test |
| 29 | 30600BZX0008 6000 | 1st ed., Mar 2025 | Analyzes breast ultrasound images to help detect suspicious lesions and assess the need for further examination. | US | Smart Opinion, Inc. | Smart Opinion, Inc. | Standalone + Physician-assisted test |
| 18 | 30200BZX0015 0000 | 10th ed., Oct 2024 | Analyzes chest CT scans to help doctors detect and mark potential lung nodules. | CT | FUJIFILM Corporation | FUJIFILM Corporation | Standalone + Physician-assisted test |
| 31 | 30100BZX0026 3000 | 4th ed., Nov 2022 | Searches a hospital's database for past cases with imaging findings similar to a user-selected area. | Undefined | FUJIFILM Corporation | FUJIFILM Corporation | Standalone test |
| 19 | 30300BZX0024 4000 | 9th ed., Oct 2024 | Analyzes chest CT scans to help doctors find and mark potential rib fractures. | CT | FUJIFILM Corporation | FUJIFILM Corporation | Standalone + Physician-assisted test |

### Transparency in Clinical Setting Reporting

Analysis of data availability revealed significant gaps in the reporting of clinical study characteristics (Figure 2) (Table 2). In standalone performance studies, none of the products (20/20, 100%) provided complete demographic information—including nationality/ethnicity/race, sex, and age. Most products (16/20, 80%) reported only the number of cases without any demographic breakdown.

**Table 2:**
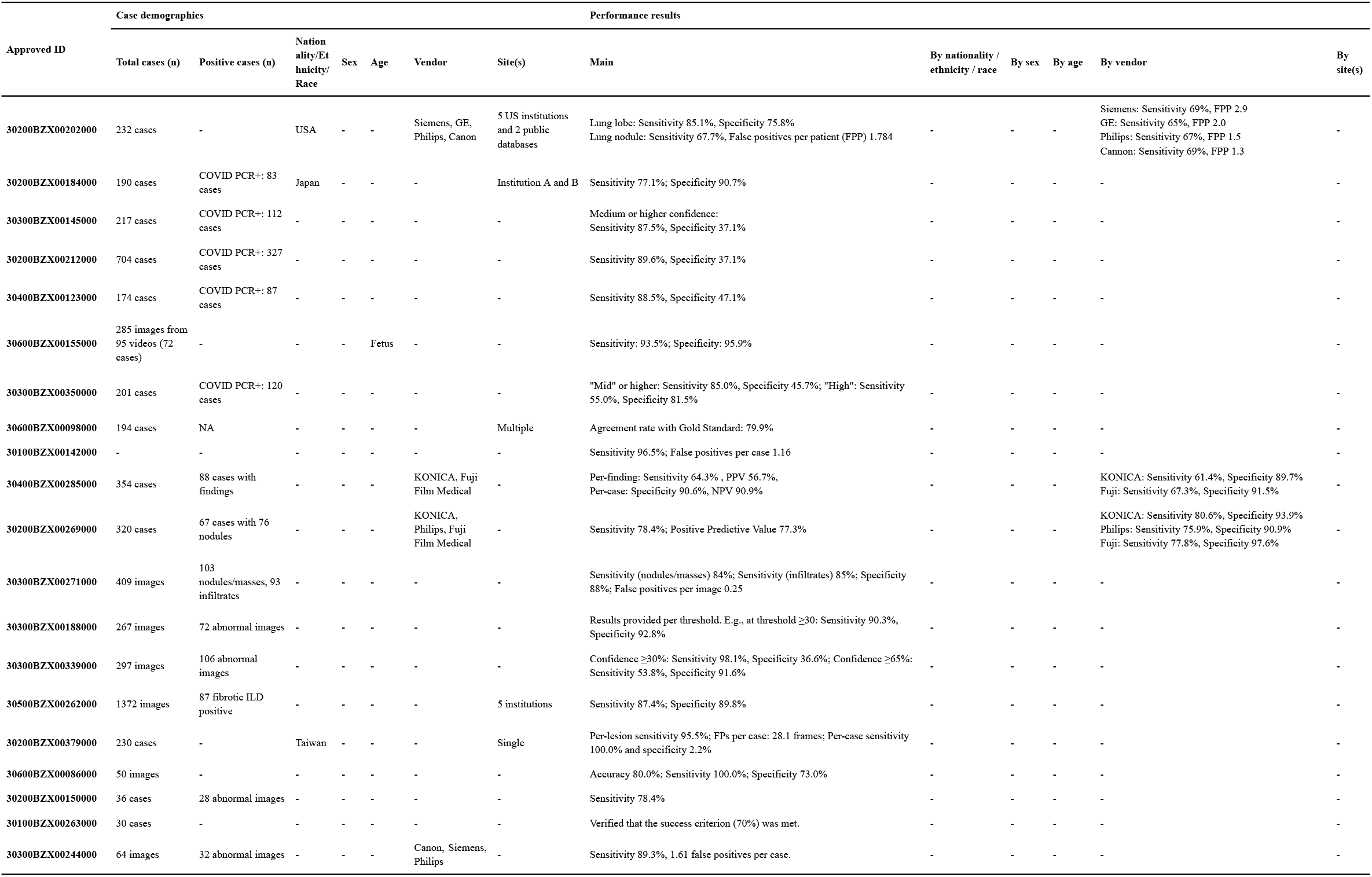
Stand-alone performance study.

**Figure 2:**
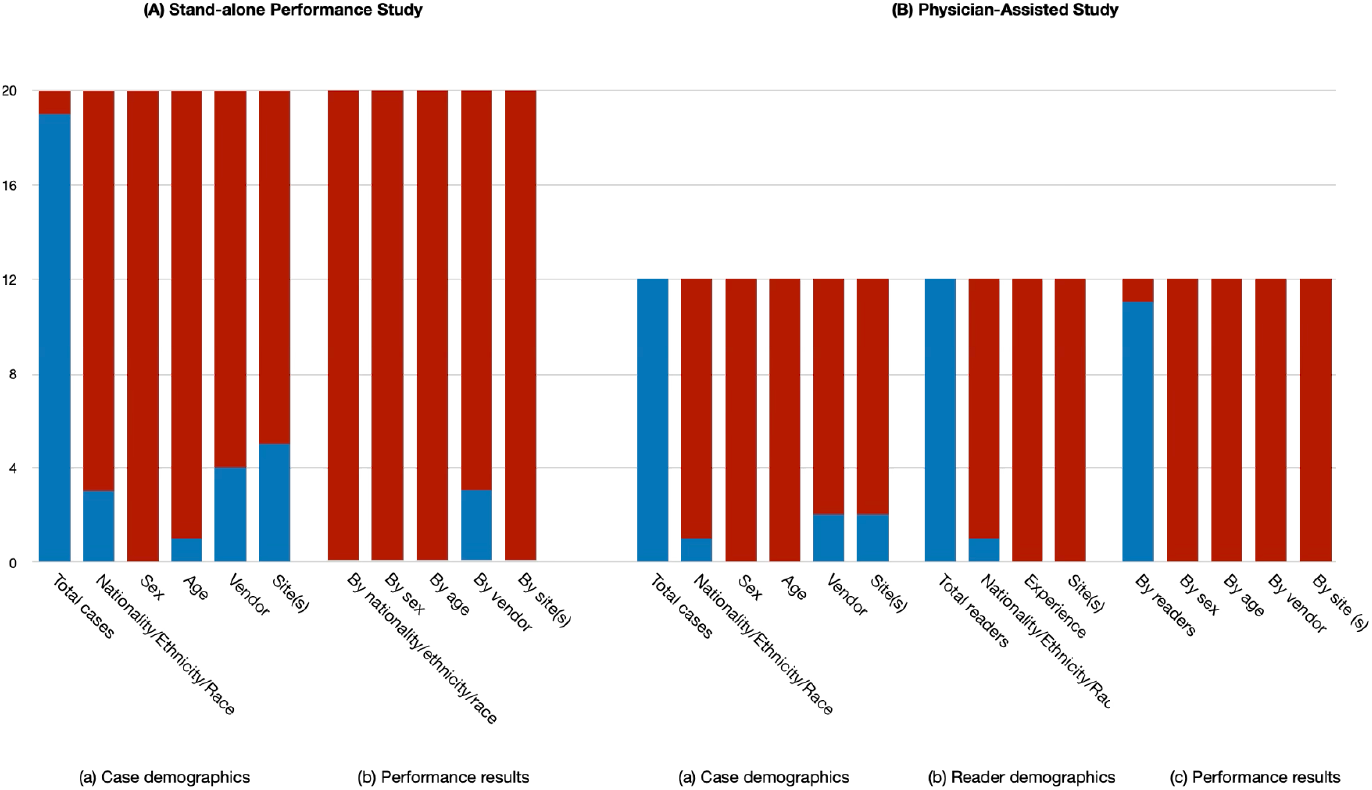
Data availability within each product. Availability of demographic and performance data for the 20 included products. The bar charts show the number of products with data publicly available in regulatory documents (blue) or not reported (red). Data availability is shown separately for (A) Stand-alone performance studies and (B) Physician-assisted studies. Categories assessed include Case demographics, Reader demographics, and Performance results stratified by various factors.

Physician-assisted studies showed similarly limited transparency in reader characteristics (Figure 2) (Table 3). Among the 12 products that included physician-assisted validation, most studies (11/12, 92%) detailed reader experience levels, but only one (1/12, 8%) reported reader nationality or practice location.

**Table 3:** Physician-assisted study.

| Approved ID | Case demographics |  |  |  |  |  |  | Reader demographics |  |  |  | Performance results |  |  |  |  |  |
| --- | --- | --- | --- | --- | --- | --- | --- | --- | --- | --- | --- | --- | --- | --- | --- | --- | --- |
|  | Total cases (n) | Positive cases (n) | Nationality /Ethnicity/ Race | Sex | Age | Vendor | Site(s) | Total readers (n) | Nationality/ Race | Experience | Site(s) | Main | By readers | By sex | By age | By vendor | By site (s) |
| 30200BZX00202000 | 232 cases | - | US | - | - | Siemens , GE, Philips, Cannon | Multi | 20 | US | Radiologists | - | AUC without AI: 0.744, with AI: 0.785-0.788 (p<0.001) | - | - | - | - | - |
| 30600BZX00155000 | 60 videos | 20 videos | - | - | - | - | - | 44 | - | 30 specialists<br>14 residents | - | Sensitivity without AI: 73.6%, with AI: 78.4% (p=0.005)<br>Specificity without AI: 79.1%, with AI: 86.5% (p<0.001) | Residents:<br>Sensitivity 63.2% to 68.6%<br>Specificity 79.8% to 89.3%<br>Specialist:<br>Sensitivity 78.5% to 83.0%<br>Specificity 78.8% to 85.3% | - | - | - | - |
| 30100BZX00142000 | 200 cases | 50 aneurysms | - | - | - | - | - | 20 | - | 10 radiologists<br>10 neurosurgeon | - | FOM without AI: 0.7168, with AI: 0.7514 (p<0.001) | Radiologists <5 years exp: 0.6470 to 0.6972<br>Radiologists ≥5 years exp: 0.7990 to 0.8218<br>Neurosurgeon <6 years exp: 0.6445 to 0.6813<br>Neurosurgeon ≥6 years exp: 0.8096 to 0.8362 | - | - | - | - |
| 30400BZX00285000 | 354 cases | 88 cases with findings | - | - | - | - | - | 10 | - | 3 radiologists<br>7 non-specialists | - | FOM without AI: 0.714, with AI: 0.771 (p<0.001) | Radiologists: 0.713 to 0.772<br>Non-specialists: 0.714 to 0.771 | - | - | - | - |
| 30200BZX00269000 | 320 cases | 67 cases with 76 nodules | - | - | - | - | - | 18 | - | 9 radiologists<br>9 non-specialists | - | AUC without AI: 0.7088, with AI: 0.7688 (p<0.001) | Radiologists: 0.7173 to 0.7683<br>Non-specialists: 0.7002 to 0.7693 | - | - | - | - |
| 30300BZX00271000 | 200 cases | 60 with findings | - | - | - | - | - | 12 | - | 4 radiologists<br>8 internal physicians | - | FOM without AI: 0.756, with AI: 0.878 (p<0.025) | Physicians ≤5 years exp: 0.751 to 0.870<br>Physicians ≥6 years exp: 0.761 to 0.887 | - | - | - | - |
| 30300BZX00188000 | 267 images | 72 abnormal images | - | - | - | - | - | 10 | - | 3 specialists<br>7 non-specialists | - | FOM without AI: 0.771, with AI: 0.839 (p<0.05) | Specialists: 0.837 to 0.862<br>Non-specialists ≥10 years exp: 0.724 to 0.829<br>Non-specialists <5 years exp: 0.756 to 0.829 | - | - | - | - |
| 30500BZX00262000 | 85 images | 19 fibrotic ILD | - | - | - | - | Multi | 25 | - | 5 specialists<br>20 non-specialists | - | AUC without AI: 0.8079, with AI 0.8457 (p<0.0001) | Specialists: 0.8917 to 0.8969 (p=0.1181)<br>Non-specialists: 0.7876 to 0.8313 (p=0.0002) | - | - | - | - |
| 30200BZX00379000 | 51 cases | - | - | - | - | - | - | 18 | - | 6 specialists<br>6 junior doctors<br>6 medical technologist | - | FOM without AI: 0.6304, with AI: 0.7726 (p < 0.0001) | Specialists: 0.6245 to 0.7430<br>Junior doctors: 0.5837 to 0.7706<br>Medical technologist: 0.6830 to 0.8042 | - | - | - | - |
| 30600BZX00086000 | 50 images | - | - | - | - | - | - | 24 | - | 4 specialists<br>20 non-specialists | - | Accuracy without AI: 69.3%, with AI: 73.1% (p=0.005) | Specialists: 75.5% to 76.5%<br>Experienced non-specialists: 77.1% to 79.6%<br>Unexperienced non-specialists: 64.7% to 69.2% | - | - | - | - |
| 30200BZX00150000 | 36 images | 28 abnormal | - | - | - | - | - | 10 | - | 5 specialists<br>5 residents | - | FOM without AI: 0.638, with AI: 0.674 (p<0.05) | Specialists: 0.668 to 0.702<br>Residents: 0.609 to 0.646 | - | - | - | - |
| 30300BZX00244000 | 64 images | 32 abnormal images | - | - | - | Canon, Siemens , Philips | - | 10 | - | 5 specialists<br>5 residents | - | FOM without AI: 0.782, with AI: 0.820 (p<0.01) | Specialists: 0.765 to 0.821<br>Residents: 0.799 to 0.820 | - | - | - | - |
| AUC: Area Under the Receiver Operating Characteristic curve, FOM: Figure of Merit |  |  |  |  |  |  |  |  |  |  |  |  |  |  |  |  |  |

### Clinical Performance Metrics

Standalone performance studies demonstrated wide variability in reported metrics (Table 2). Differences in experimental design, reported outcomes, and chosen evaluation metrics meant that comparability across products was not necessarily high. Sensitivity values ranged from 55.0% to 100%, with COVID-19 detection applications generally showing variable performance depending on confidence thresholds. Specificity showed even greater variability, ranging from 2.2% to 97.6%, with several products reporting specificity below 50%.

Physician-assisted studies consistently demonstrated performance improvements with AI assistance (Table 3). Twelve products reporting physician-assisted validation showed statistically significant improvements in primary performance metrics. The magnitude of improvement varied considerably across studies. Notably, less experienced readers showed greater performance gains, with one study reporting AUC improvement from 0.6470 to 0.6972 for junior neurosurgeons compared to 0.7990 to 0.8218 for experienced radiologists [30].

### Subgroup Performance Analysis

Critical gaps were identified in subgroup performance reporting. No products provided performance metrics stratified by patient age, sex, or ethnicity, despite the known impact of demographic factors on AI performance. Only 3 products (15.0%) reported vendor-specific performance differences, revealing substantial variations. For example, one lung nodule detection system showed sensitivity ranging from 65% on GE equipment to 69% on Canon systems, with false positive rates varying from 1.3 to 2.0 per patient across vendors. Site-specific performance data was absent from all package inserts, preventing assessment of generalization across different clinical settings.

## Discussion

This scoping review reveals fundamental gaps in the transparency of clinical evidence for PMDA-approved AI-based radiology software, with significant implications for clinical implementation and patient safety. The finding that none of the products provide complete demographic information and none offer subgroup-stratified performance metrics represents a critical barrier to evidence-based adoption of these technologies in Japanese healthcare settings.

### Implications for Clinical Implementation

The wide variability in performance metrics underscores the importance of understanding the specific clinical context and validation conditions for each AI system [4]. Without detailed demographic information or site-specific performance data, clinicians cannot adequately assess whether published performance metrics will translate to their specific patient populations. This uncertainty may lead to either inappropriate rejection of beneficial technologies or, conversely, over-reliance on systems that may not perform as expected in local contexts [9].

Of the twenty AI-based radiology products, twelve included physician-assisted studies. The consistent performance improvements demonstrated in physician-assisted studies suggest that AI integration can enhance diagnostic accuracy across experience levels. However, the greater gains observed among less experienced readers raise important questions about training requirements and the potential for AI to either reduce or exacerbate expertise-related disparities in diagnostic performance [32]. No physician-assisted study reported performance results other than by-reader analyses, a notable omission given the heterogeneity of patient populations, imaging protocols, and clinical workflows across Japanese healthcare institutions.

The five COVID-19 detection applications warrant separate analysis given their rapid development and deployment timeline, reflecting the pandemic’s influence on AI development priorities. All of these products were approved based solely on standalone performance studies without reader studies. Consequently, each product received conditional approval requiring that appropriate post-marketing studies be conducted to evaluate the product’s performance and clinical utility. These products showed variable performance metrics, with sensitivity ranging from 55.0% to 89.6% (depending on confidence thresholds) and specificity from 37.1% to 90.7%. None provided information about variant-specific performance or temporal validation despite the evolving nature of COVID-19 imaging findings.

### Regulatory Evolution and International Comparisons

Japan’s PMDA has been progressive in establishing approval pathways for AI-based medical devices, yet our findings suggest that documentation requirements have not evolved to match the unique characteristics of AI systems [33]. International regulatory bodies, including the Food and Drug Administration and European authorities, have begun implementing more stringent requirements for AI transparency, including mandatory reporting of training data characteristics and performance across demographic subgroups [5]. The absence of such requirements in current PMDA package inserts places Japanese healthcare providers at a disadvantage in making informed implementation decisions.

The contrast between Japan’s advanced regulatory framework for AI approval and the limited transparency requirements for clinical evidence creates a paradox. While the PMDA’s efficiency in approving AI products has positioned Japan in the top tier of medical AI adoption, the lack of detailed performance information may ultimately hinder successful clinical integration and potentially compromise patient safety [3].

### The Role of Real-world Performance Monitoring

The gaps identified in pre-market clinical evidence underscore the critical importance of post-market surveillance and real-world performance monitoring for AI-based medical devices. Unlike traditional medical devices, AI systems can exhibit performance drift over time due to changes in patient populations, imaging protocols, or clinical practices [34]. Current package insert requirements do not address this dynamic nature of AI performance, nor do they provide frameworks for ongoing validation.

This situation creates opportunities for specialized organizations that can bridge the gap between regulatory compliance and clinical implementation needs. Companies with expertise in Japan’s Next Generation Medical Infrastructure Act and Personal Information Protection Act are uniquely positioned to facilitate privacy-preserving, multi-institutional validation studies that can provide the real-world evidence lacking in current regulatory documentation. Such organizations can establish standardized frameworks for continuous performance monitoring, ensuring that AI systems maintain their expected performance across diverse clinical settings while complying with Japan’s strict data privacy regulations.

### Recommendations for Enhanced Transparency

Based on our findings, we propose several recommendations for enhancing transparency in AI-based radiology software documentation. First, package inserts should include mandatory reporting of demographic characteristics for both training and validation datasets, with performance metrics stratified by key demographic variables. Second, vendor-specific and site-specific performance data should be required when technically feasible, acknowledging the impact of imaging equipment and clinical workflows on AI performance.

Third, for physician-assisted studies, detailed reader characteristics including specialization, experience levels, and geographic distribution should be standard requirements. This information is essential for healthcare institutions to assess the training and implementation requirements for their specific workforce. Fourth, products should include clear statements about the reference standards used for validation, particularly important for conditions like COVID-19 where diagnostic criteria have evolved over time.

### Industry Innovation and Future Directions

The current regulatory landscape creates both challenges and opportunities for innovation in medical AI validation and implementation. Organizations specializing in medical data governance and AI validation can play a crucial role in establishing industry standards that exceed minimum regulatory requirements. By leveraging frameworks such as the Next Generation Medical Infrastructure Act, these organizations can create secure, privacy-preserving environments for continuous AI validation across multiple institutions.

Furthermore, the development of standardized reporting templates and performance monitoring dashboards could significantly enhance transparency while reducing the burden on individual manufacturers. Industry-led initiatives for voluntary transparency standards, potentially coordinated through professional societies or industry associations, could accelerate the evolution of regulatory requirements while immediately benefiting clinical users.

### Limitations

First, our analysis was limited to publicly available package inserts; additional information accessible through manufacturer communications or training materials could not be captured. Second, we lacked access to the raw clinical-study data underlying package-insert claims, preventing independent verification of performance metrics and deeper assessment of study quality. Third, this review focused exclusively on PMDA-approved products; third-party–certified AI-SaMD, which are more numerous in Japan, were not evaluated and may display different transparency patterns.

### Conclusion

This scoping review of PMDA-approved AI-based radiology software reveals significant gaps in the transparency and completeness of clinical evidence available to healthcare providers. The absence of demographic stratification, vendor-specific performance data, and detailed validation conditions creates uncertainty for clinicians and healthcare institutions seeking to adopt these technologies. Addressing these gaps requires coordinated efforts from regulators, manufacturers, and specialized organizations with expertise in medical data governance and AI validation. Enhanced transparency standards, coupled with robust frameworks for continuous real-world performance monitoring, are essential to unlock the full potential of AI in Japanese radiology practice while safeguarding clinical effectiveness and patient safety.

## Data Availability

All data used in this study are publicly available regulatory documents provided by the Pharmaceuticals and Medical Devices Agency (PMDA), Japan. No additional datasets were generated or analyzed.

